# Leveraging Large Language Models for Identifying Interpretable Linguistic Markers and Enhancing Alzheimer’s Disease Diagnostics

**DOI:** 10.1101/2024.08.22.24312463

**Authors:** Tingyu Mo, Jacqueline C. K. Lam, Victor O. K. Li, Lawrence Y. L. Cheung

## Abstract

Alzheimer’s Disease (AD) is a progressive irreversible neurodegenerative disorder. Early AD detection is crucial for timely intervention. This study proposes a novel LLM framework to identify interpretable linguistic markers from LLMs and incorporate them to supervised AD detection transformers, while evaluating corresponding model performance and interpretability. Our work carries three major novelties: First, we design in-context Few-Shot and Zero-Shot prompting strategies to facilitate LLMs in summarising the high-level linguistic markers discriminative of AD from Normal Control (NC), providing interpretation and assessment of their strength, reliability and relevance for AD classification. Second, we incorporate LLM-summarised linguistic markers into a smaller transformer model to enhance the performance of AD detection. Third, we investigate whether the LLM-summarised linguistic markers can enhance accuracy and interpretability of our downstream supervised transformer model when used with the original speech transcripts. Our findings have shown that using LLM-summarised linguistic markers solely may yield lower accuracy using supervised learning, while combining these markers with the features extracted from the original speech transcripts can enhance the AD detection model’s diagnostic capabilities. Our hybrid speech-based AD detection framework capitalizes both the interpretability of linguistic markers and the rich information embedded within the original transcripts. When used as complementary features, the distinguishable AD linguistic markers have enhanced the AD detection performance, while providing clinically meaningful insights into speech-based AD detection. These insights have enabled healthcare professionals to gain deeper understandings about what linguistic variations have dictated individuals suffering from AD, enabling the clinical professionals to make more informed interventions and healthcare decision-makings.

## 1 Introduction

Alzheimer’s Disease is a devastating neurodegenerative disorder that progressively impairs cognitive function, affecting millions of individuals worldwide. Timely and precise diagnosis of AD is of utmost importance, as it enables early therapeutic intervention and optimal healthcare management, potentially improving patient outcomes and their qualities of life. Recent advancements in natural language processing (NLP) ^24^,^26^and deep learning offer new opportunities for developing automated tools to assist the detection and diagnosis of AD ^1,2,27,28^NLP models, such as BERT (Bidirectional Encoder Representations from Transformers), ^4^ have achieved remarkable performance in various NLP tasks, including language understanding, generation and classification. However, their applications in healthcare contexts demand careful balance between model performance and interpretability: First, it is necessary for clinicians to interpret and validate diagnostic model findings against clinical decisions, based on a conventional set of clinical indicators. Second, accurate early-stage AD detection is dependent on clear, interpretable clinical biomarkers that discriminate cognitive decline from normal aging. Third, healthcare providers must be able to communicate findings effectively with patients, particularly when it is suitable for early intervention. While more complex NLP models might detect subtle linguistic patterns at a higher accuracy, their “black box” nature may limit clinical adoption. Conversely, more interpretable models might sacrifice statistical performance, though they may offer clearer insights for clinical validation, thus enhancing the potential of integrating the model results with the process of clinical diagnostics.

Traditional NLP methods for AD detection can achieve reasonable accuracy in distinguishing AD from NC. Automated speech analyses ^11^ have shown promising results in predicting AD progression, achieving an accuracy of 78.5%. However, they are weak in capturing /interpreting the underlying linguistic patterns. The complex nature of linguistic change in AD requires more sophisticated models that not only detects linguistic variations but also articulates them in meaningful ways that are intelligible by the healthcare professionals. Additionally, the limited availability of annotated speech data from individuals suffering from AD pose a substantial challenge for conventional supervised learning models operated on large datasets. Large Language models (LLMs) offer a promising direction for addressing these challenges through Few-Shot and Zero-Shot in-context learning, capable of learning based on limited data size/samples. LLMs are deep learning models trained on vast amounts of textual data, enabling them to capture rich linguistic information by retrieving domain-specific knowledge from big clinical datasets. More importantly, they are capable of identifying and interpreting high-level linguistic patterns which may be used as interpretable indicators of cognitive decline, complementing existing diagnostic biomarkers. Interpretability is essential for integrating model results with clinical practice, as healthcare providers need clear, explainable evidence to support clinical diagnostic decisions. For instance, interpretable linguistic biomarkers of AD identified from interpretable LLM models, such as repetition and coherence, can potentially serve as clinical linguistic biomarkers to assist clinicians in making more informed AD diagnostics.

In this paper, we propose a novel approach for AD detection based on interpretable linguistic biomarkers summarised from LLMs. Our method leverages Few-Shot and Zero-Shot learning techniques to enable LLMs to identify high-level meaningful linguistic patterns from limited transcription data. We design a set of prompting strategies to guide LLMs in summarising key linguistic features and generate explanations. Furthermore, we incorporate LLM-summarised linguistic biomarkers into LLMs for downstream supervised learning, with the final goal of assessing AD versus NC classification performance (for AD detection). While LLMs have achieved impressive results in various NLP performance tasks, their AD classification performance is still lagging behind traditional supervised learning approaches, given the limited data constraint. To overcome this challenge, we propose a novel AD detection architecture that capitalizes on the strengths of both LLMs and supervised learning techniques. Our main contributions and novelties are as follows:

- Use Large Language Models (LLMs) to extract interpretable linguistic markers for AD vs. NC classification, or AD detection, using in-context Few-Shot and Zero-Shot prompting strategies.
- Design different prompting strategies under different learning contexts to facilitate LLMs in identifying high-level linguistic markers discriminative of AD/NC, providing interpretation and assessment, in terms of their strength, reliability and relevance to AD vs. NC classification, or AD detection.
- Incorporate linguistic markers summarised by LLMs into a supervised-learning model to enhance the performance of downstream supervised learning for AD vs. NC classification, or AD detection, by fusing high-level linguistic markers/features summarised from LLMs with the original speech transcript.

The remainder of this paper is organized as follows. Section 2 provides an overview of the related work covering AD classification/detection using NLP and deep learning techniques. Section 3 describes the methodology, including prompt engineering strategies, and outlines our proposed model framework for evaluation. Section 4 presents the experimental results and discusses the performance of our proposed model. Finally, Section 5 and Section 6 concludes the paper and outlines the potential for future studies.

## 2 Related Work

Recently, there has been a growing interest in using linguistic biomarkers in AD diagnostics. Linguistic markers refer to the distinctive linguistic features and patterns that are indicative of cognitive decline. Several studies have explored the potential of linguistic markers in AD detection. Fraser et al. ^5^ analyzed a wide range of linguistic features, including grammatical complexity, semantic content, and discourse coherence, in the language samples of AD patients and healthy controls. They found that a combination of linguistic features could effectively distinguish between AD and normal aging. Similarly, Orimaye et al. ^6^ used natural language processing techniques to extract syntactic, lexical, and semantic features from the transcripts of AD patients and achieved promising results in AD classification. Other studies have focused on specific linguistic aspects, such as speech fluency, word-finding difficulties, and semantic fluency. For example, Pakhomov et al. ^19^ investigated the use of verbal fluency tests as a screening tool for frontotemporal lobar degeneration, while demonstrating their effectiveness in detecting cognitive impairment. Kavé and Goral ^7^ explored the semantic and phonemic fluency deficits in AD and highlighted their potentials to serve as linguistic markers of AD.

In the medical domain, LLMs have been used for tasks such as medical language translation, clinical note summarization, and medical question answering. For example, Rasmy et al. ^10^ developed a medical language model called MED-BERT, which was trained on a large corpus of medical text and achieved state-of-the-art performance on several medical natural language processing tasks. Some studies have explored the potential of LLMs in mental health analysis. Yang et al. ^13^,^14^ proposed interpretable mental health analysis frameworks using LLMs to identify linguistic patterns and generate explanations from text data. Xu et al. ^15^ introduced ExpertPrompting, instructing LLMs to behave as domain experts, enhancing performance and reliability in mental health applications. Luo et al. ^16^ explored ChatGPT’s capability in evaluating factual inconsistencies, highlighting the potential of LLMs in assessing the reliability of generated insights. Jeon et al. ^17^ proposed a dual-prompting approach to improve the interpretability and reliability of mental health analysis using LLMs. These studies have demonstrated the growing interest in leveraging LLMs for accurate, interpretable, and trustworthy mental health analyses. Our research builds upon these advancements, exploring the use of LLMs for identifying interpretable linguistic markers in Alzheimer’s Disease detection. We evaluate the feasibility of using LLMs to identify interpretable linguistic markers for detection of AD based on Few-Shot and Zero-Shot learning. We design various prompting strategies under different learning contexts to facilitate LLMs in identifying linguistic biomarkers discriminative of AD, assessing their strength and reliability, and providing interpretation concerning their relevance to AD detection. We use novel linguistic biomarkers summarised by LLMs to enhance the performance of downstream supervised learning, by proposing a novel framework that incorporates the LLM-identified markers into a multimodal AD detection model. Our work contributes significantly to the growing body of literature covering LLM-assisted AD diagnostics.

## 3 Methodology

We propose a feasible approach to evaluate linguistic markers identified by LLMs for Alzheimer’s disease (AD) detection. We design two prompting strategies that leverage the knowledge and capabilities of LLMs to analyze the linguistic patterns that govern individual speeches.

### 3.1 LLM Configuration for Model Development

We explore the capabilities and applications of several leading LLMs, including ChatGPT3.5, GPT4, GPT4o, to extract linguistic patterns and markers relevant to Alzheimer’s disease (AD) diagnosis. These models are representatives in LLM technology, each having unique characteristics and strengths in generating and analyzing human languages.

- **GPT3.5**: As a version of GPT (Generative Pre-trained Transformer) series developed by OpenAI, it serves as one of the most powerful language models available during the time of its release. It excels in generating human-like text, completing given text prompts at high coherence, and understanding subtle linguistic nuances. We leverage GPT-3.5’s capability to identify the linguistic patterns and anomalies indicative of cognitive impairment or AD.
- **GPT4/4o**: GPT-4/4o is the more sophisticated version fine-tuned for better understanding and generating natural language. While specific details regarding its size and architecture are proprietary, it is known for its enhanced ability to process and generate text across a wide range of languages and dialects, providing more accurate and contextually relevant responses. While GPT-4 excels at text processing and generation, GPT-4o is designed for comprehensive multimodal interaction, accepting inputs in text, audio, image, and video formats while generating outputs in text, audio, and image modalities. We evaluate GPT4/4o’s advanced language understanding to extract more refined and contextually-aware linguistic markers.

### 3.2 LLM-based Prompting Strategies for Linguistic Markers Identification

We propose a novel approach to identify linguistic markers for Alzheimer’s Disease (AD) detection using LLMs. We design a prompting strategy that leverages the knowledge and capabilities of LLMs to analyze linguistic patterns in descriptions of the Cookie Theft picture from the Boston Diagnostic Aphasia Examination. The methodology is outlined in Algorithm 1.

As shown in Line 3-4, the prompting process begins by assigning an expert identity ^15^ to the LLM, portraying it as a medical professional tasked with analyzing linguistic patterns in the Cookie Theft picture descriptions. This identity establishment helps to guide the LLM’s behavior and outputs towards the desired task. Next, we assign the specific task to the LLM, which is to identify linguistic indicators that distinguish between individuals with AD and those with normal control. We emphasize the importance of being cautious and avoiding overdiagnosis of AD based solely on linguistic patterns. We use two prompting strategies to enable LLM’s potential abilities, including Zero-Shot prompting and Few-Shot prompting.

- **Zero-Shot Prompting**: Zero-Shot learning ^9^ enables LLMs to apply learned knowledge from one domain to perform tasks in another without any task-specific training data. We aim to prompt the general LLMs to describe linguistic attributes found in transcripts, then relate these attributes to known symptoms or stages of AD without explicit example-based guidance. Zero-Shot prompting strategy includes two approaches: a. Inferential Question Prompt: This kind of prompt directly asks the LLM to infer cognitive decline based on the language patterns observed in the transcript. For example, “What language patterns in this transcript suggest cognitive decline?” This prompt encourages the LLM to identify and analyze linguistic features that may indicate cognitive impairment. b. Direct Inquiry Prompt: This prompt explicitly instructs the LLM to identify any indicators of cognitive impairment in the given transcript. For example, “Identify any cognitive impairment indicators in the following transcript.” This prompt directs the LLM’s attention towards detecting specific linguistic markers associated with cognitive decline. In our method, we simply adopt Direct Inquiry Prompt to instruct the LLM, as shown in Line 11 in Algorithm.1 as follows:

> *“Identify linguistic patterns, keywords, or phrases that could potentially indicate cognitive impairment or Alzheimer’s disease. However, be cautious and consider whether these indicators are strong enough to warrant a diagnosis on their own. Present these indicators and your assessment of their strengths in a list format.”*
- **Few-Shot Prompting**: Few-Shot learning ^8^ involves training the LLM models with extremely limited labelled data. We aim to fine-tune the LLMs with AD-specific linguistic markers using limited data from transcripts. We design a general LLM model without specifying any specific domains using Few-Shot prompting. Few-Shot prompting is conducted using extremely limited speech transcript data collected from AD and NC cohorts, followed by a question that seeks to identify similar linguistic patterns in the much larger dataset employed for LLM model training. Few-Shot prompting is conducted based on the following Chain-of-Thoughts sequences:

> *“<Examples> Transcripts from people with Alzheimer’s Disease describing the Cookie Theft picture: Script: [example1]*…*[exampleK]*
>
> *<Examples> Transcripts from people who are normal control describing the Cookie Theft picture: Script: [example1]*…*[exampleK]”*.

In this prompting sequence, the LLM model is provided with a small number of example transcripts (K examples) indicating the presence or absence of a specific condition (e.g., AD or NC). By employing the Zero-Shot and Few-Shot prompting strategies, we aim to evaluate whether LLMs can leverage domain-specific knowledge and extract linguistic markers that discriminate AD from NC. The summarised linguistic markers will be utilized in downstream supervised learning for examining AD detection performance.

### 3.3 LLM-based Interpretable Explanation Generation

To analyse further the LLM-identified linguistic markers and enhance model interpretability, we assign two novelites/tasks for the LLM-based AD detection model. As shown in Line 13 in Algorithm 1, Task (b) focuses on making a diagnostic decision based on the LLM-identified linguistic indicators and their AD classification strength. The prompt is given as follows:

> *“Based on the identified linguistic indicators and their strength, make a diagnosis decision (Alzheimer’s Disease or normal control) for the individual who provided the description. If the indicators are not strong or conclusive enough, lean towards a diagnosis of Not Sure”*.

We instruct the LLM to classify the individuals who provided the Cookie Theft picture description as either suffering from Alzheimer’s Disease (AD) or cognitively normal, which we termed as Normal Control (NC). However, we emphasize the importance of caution in this decision-making process. If the LLM-identified linguistic indicators are not strong or conclusive enough to confidently determine whether an individual has suffered from AD, we guide the LLM to lean towards a decision of “Not Sure.” This approach aims to prevent over-diagnosis and ensures that the LLM has taken into account the reliability and significance of linguistic markers before making any definitive AD classification.

As shown in Line 14 in Algorithm.1, Task (c) involves generating a concise explanation for the diagnosis decision made in Task (b). Our prompt for this task is as follows:

> *Generate a concise explanation for your diagnosis decision. Discuss how the identified keywords, phrases, and patterns from the transcript support your conclusion, and explain any reservations you have about the strength of the indicators, while emphasizing the importance*.

We prompt the LLM to discuss how the identified keywords, phrases, and patterns from the transcript support its conclusion. The LLM-based explanation should highlight the specific linguistic features that contribute to the classification decision, providing insights into the reasoning process of the LLM. Additionally, we instruct the LLM to provide any reservations or uncertainties regarding the strength of the linguistic indicators. By doing so, we encourage the LLM to critically assess the reliability and capture the richness and complexity of the linguistic markers, while disclosing the limitations or ambiguities in the concerned LLM-based decision-making process. This step highlights the cautious approach we have taken in model design to avoid over-diagnosis, while promoting transparency and interpretability of model results.

#### Algorithm 1 Prompting LLMs for Analysing Linguistic Patterns in Cookie Theft Picture Descriptions

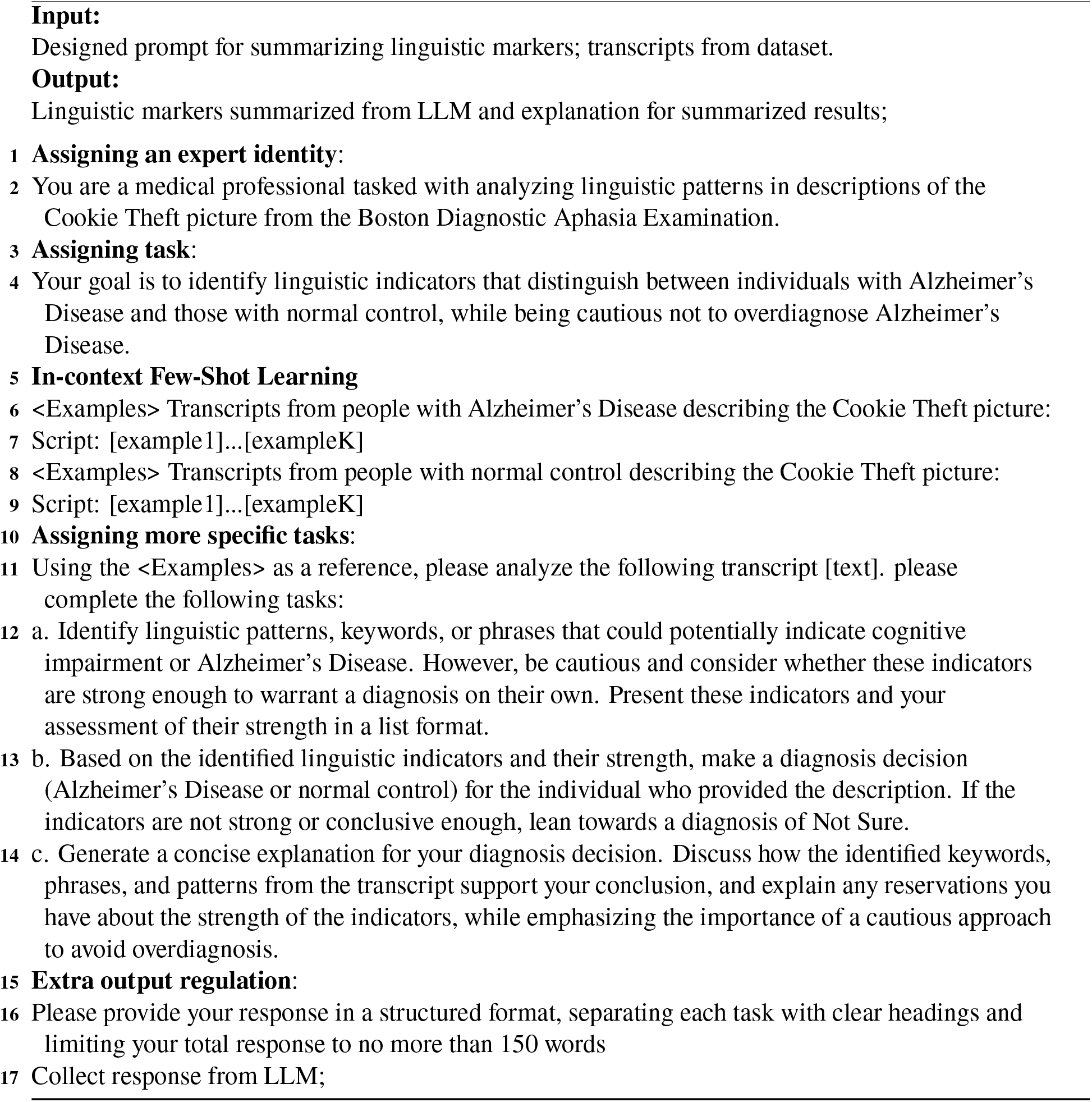

### 3.4 Evaluating Linguistic Markers in AD Diagnostic Framework

We propose an evaluation framework to assess the effectiveness and interpretability of the linguistic markers summarised by the LLMs for AD detection. We aim to investigate whether these linguistic markers can replace the original transcripts in training AD detection models, while evaluating the performance of different prompting strategies and interpretability of our LLMs. We train the AD diagnostic models, using the LLM-identified linguistic markers and interpretations as the input features, then compare their performance to other models trained on the original speech transcript data. We employ various evaluation metrics, including, accuracy, precision, recall, and F1 score, to assess the model performance. If the models trained on the linguistic markers have achieved comparable or better performance than those trained on the original transcripts, it indicates that the summarised markers have captured the relevant information for AD detection, thereby potentially obviating the less interpretable speech transcripts.

Referring to Fig.1, our framework consists of two main components: (1) the speech embedding branch and (2) the text embedding branch. As demonstrated in, ^12^ representation learning can improve the naturalness of speech-driven gesture generation. On the one hand, the speech embedding branch in our framework processes the sampled raw speech using a speech encoder to generate speech embeddings. On the other hand, the text embedding branch utilizes the summarised linguistic markers and their corresponding interpretations. The linguistic markers serve as an alternative representation of the original speech, capturing the relevant information for AD detection. These markers are then fed into the text encoder, to be converted into the text embeddings for the subsequent processing steps, which consist of embedding fusion and classification using the Multi-layer Perceptrons (MLP) head.

**Figure 1.**
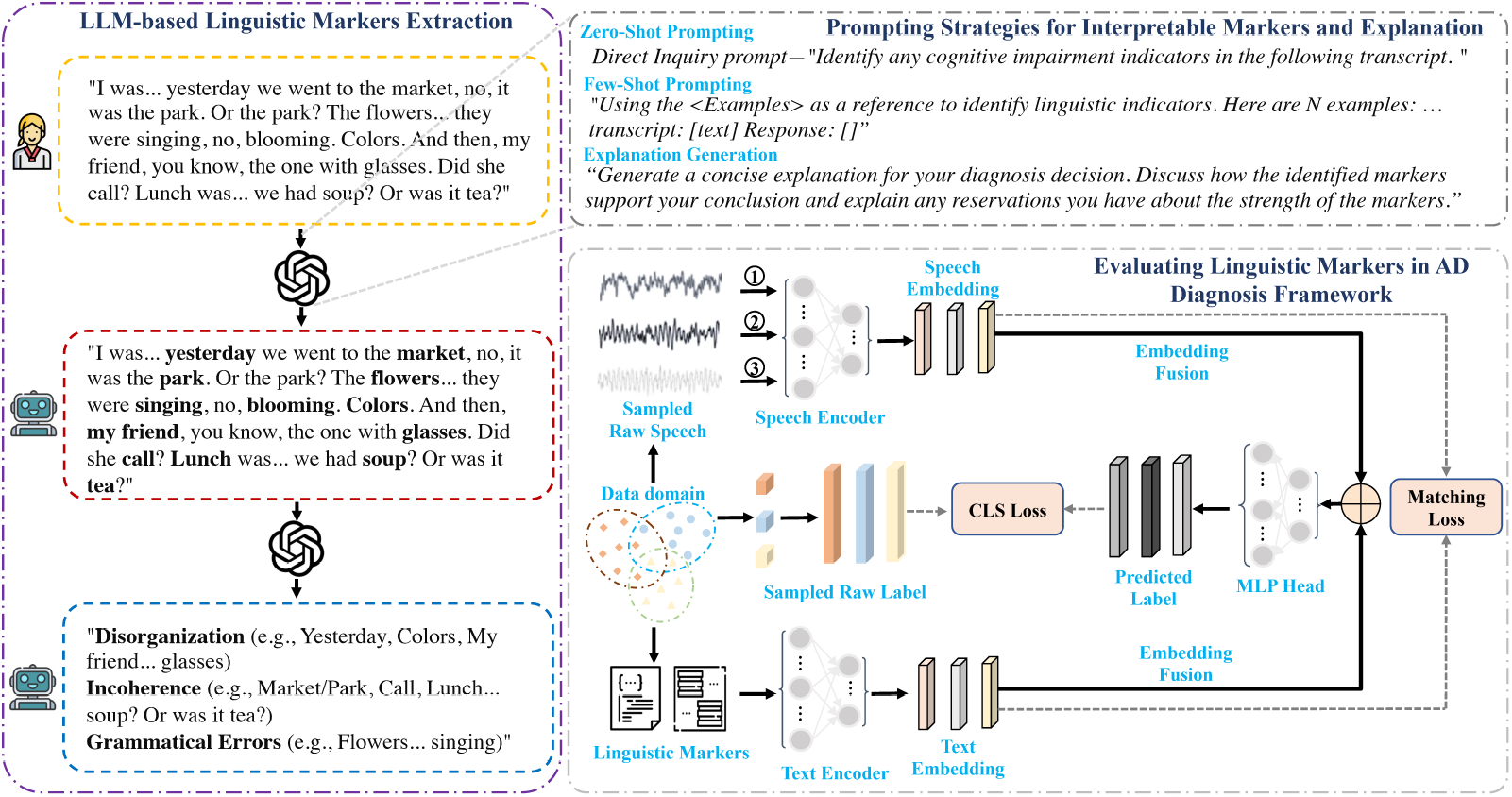
A framework for leveraging LLMs for identifying interpretable linguistic markers and enhancing Alzheimer’s Disease diagnostics.

To train the AD detection model, we employ two loss functions: the cross-entropy loss and the matching loss. The cross-entropy loss is used to measure the discrepancy between the predicted labels and the ground truth labels, guiding the model to give accurate AD diagnosis predictions. The matching loss, on the other hand, is designed to align the speech embeddings with text embeddings. By minimizing the matching loss, we encourage the model to learn a shared representation space where the speech and text embeddings are closely aligned. The optimization objects are listed in follows:

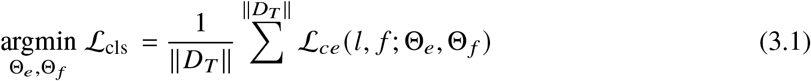

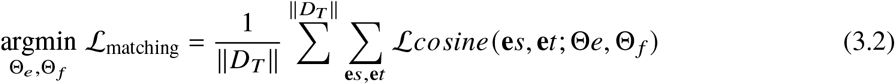

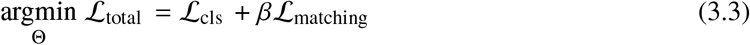

where *f* is the fused embedding, Θ_*e*_ and Θ _*f*_ are the parameters of encoders and MLP-Head for classification respectively. *ℒ*_*ce*_ represents the cross-entropy loss and *ℒ*_matching_ represents the matching loss. |*D*_*T*_ | denotes the size of the training dataset. **e***s* and **e***t* are the speech embedding and text embedding, respectively. *ℒ*_*cosine*_ is the cosine distance based loss. *β* is the weight for balancing these objects.

## 4 Experiment

### 4.1 Evaluation Datasets

We utilize the ADReSSo Challenge 2021 dataset, ^20^ the Pitt corpus ^33^ and FDRD dataset to investigate the feasibility of using LLMs for identifying interpretable linguistic markers of AD and improving the performance of AD detection. The datasets deployed consist of spontaneous speech from diverse AD-related and NC cohorts, making it a valuable resource for advancing our understanding on what linguistic markers are characterizing symptomatic AD. More specifically, the speech recordings were obtained as a result of the Cookie Theft picture description task performed by different bodies, in accordance with the Boston Diagnostic Aphasia Examination. For this task, participants are shown a picture depicting a scene where a boy is stealing cookies from a jar, while his mother is washing her dishes. The scene also depicts other objects such as a sink overflowing with water and a boy trying hard to get hold of a cookie jar located at the top part of the shelf. Participants are asked to describe what they have observed in the picture, in order to collect spontaneous speech samples based on which their language abilities and cognitive functions can be evaluated.

**Table 1.**
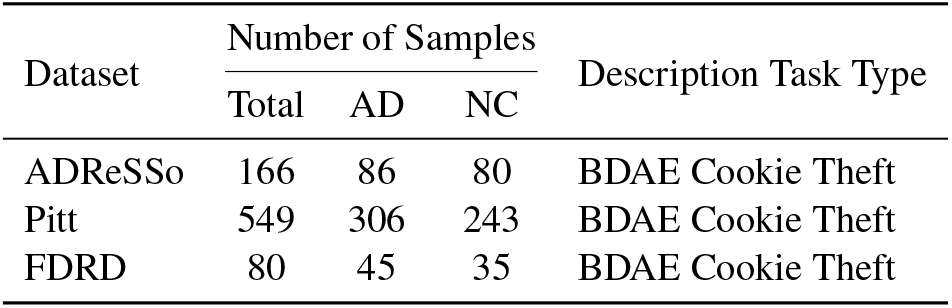
An overview of the speech datasets adopted for this study. AD: Alzheimer’s Disease; NC: Normal Control; BDAE: Boston Diagnostic Aphasia Examination.

#### 4.1.1 The ADReSSo Dataset

The ADReSSo Challenge 2021 dataset is widely used in the research community, serving as a benchmark for developing and evaluating methods for AD detection through speech analysis. These spontaneous speech samples have been divided into two main categories based on the cognitive labels: Alzheimer’s Disease (AD) and Normal Control (NC). The AD category covers speech recordings and transcripts from individuals diagnosed as Alzheimer’s Disease patients. The speech samples in this category exhibit linguistic patterns and features associated with AD. NC category includes the speech recordings and the transcripts from individuals who are cognitively normal, meaning they do not display any signs of cognitive impairment or dementia. The speech samples in this category serve as the control samples and provide a baseline for comparing the linguistic characteristics of AD individuals. In our experiment, our dataset contains a total of 166 speech recordings and transcripts, with 86 samples in the AD category and 80 samples in the NC category.

#### 4.1.2 The Pitt Corpus

The DementiaBank Pitt Corpus, part of the larger TalkBank project, offers a longitudinal perspective on cognitive decline through speech analysis. This dataset complements the ADReSSo dataset by providing a larger sample size and multiple recordings for some participants over time. Our experimental subset of the Pitt Corpus contains a total of 549 audio recordings from 291 unique participants. The audio recordings are divided into two main categories: 306 recordings from participants diagnosed with dementia and 243 recordings from cognitively normal participants. Similar to the ADReSSo dataset, participants have completed the Cookie Theft picture description task, providing consistency in the speech elicitation method across both datasets.

#### 4.1.3 The FDRD Dataset

The FDRD dataset^1^ from Cambridge Centre for Frontotemporal Dementia and Related Disorders (FDRD), a clinical database consisting of speech samples from participants suffering from various neurodegenerative conditions, as well as healthy controls. Complementary data include assessments from both baseline and follow-up visits, enabling longitudinal analysis. Participants underwent standardized picture description tasks using three validated stimuli: the Boston Diagnostic Aphasia Examination (BDAE) Cookie Theft picture, the Mini Linguistic State Examination (MLSE) beach scene, and the Western Aphasia Battery (WAB) picnic scene. The dataset covers multiple diagnostic groups, including semantic variant primary progressive aphasia (svPPA), non-fluent variant PPA (nfvPPA), logopenic variant PPA (lvPPA), mixed PPA, corticobasal syndrome (CBS), progressive supranuclear palsy (PSP), and Alzheimer’s Disease (AD). In our study, we focus on two main groups: participants with AD (n=45, including those with multi-domain dementia, lvPPA, and typical amnestic AD) and neurologically healthy controls (n=35). Comprehensive participant information was collected, including age, sex, handedness, educational background, and cognitive performance as measured by the Addenbrooke’s Cognitive Examination (ACE, scoring out of 100).

### 4.2 Experimental Set-up

We provide a detailed description of the experimental setup for evaluating our proposed methodolody. Our experiments were conducted using the Python 3.8 programming language on an Ubuntu 22.04 LTS operating system. We utilized the PyTorch 1.9 deep learning framework along with additional libraries such as Huggingface for establishing our model. The experiments were performed on a system equipped with an Intel(R) Xeon(R) Platinum 8370C CPU @ 2.80GHz and four NVIDIA GeForce RTX 4090 GPUs with 24GB of VRAM. During training, we employed the AdamW optimizer with an initial learning rate of 2e-5 and a weight decay of 0.05. The batch size was set to 4. The model was trained for a total of 100 epochs. Gradient accumulation was used with a step size of 2 to effectively increase the batch size and stabilize the training process. The coefficient weight *β* for balancing our objectives was set to 0.1. We applied a linear learning rate scheduler with a warmup parameter of 0.05 to gradually increase the learning rate during the initial training stages.

### 4.3 Evaluation Metrics

To assess the performance of our proposed method for detecting Alzheimer’s Disease using the LLM- identified linguistic markers, we employ two primary evaluation metrics: Accuracy and F1 score. Accuracy is a widely used metric that measures the overall correctness of model prediction. It is calculated as the ratio of the number of correctly classified samples to the total number of samples in the dataset. In our case, Accuracy represents the percentage of correctly identified AD and NC samples. Accuracy is computed using the following formula:

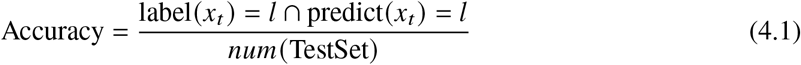

where, *x*_*t*_ is the test data, num(TestSet) is the total counts of testset samples, *label* (*x*_*t*_) is the ground truth label of *x*_*t*_ and *predict* (*x*_*t*_) is the predicted label of *x*_*t*_ by the trained classifier for AD detection. While Accuracy provides an overall measure of the model’s performance, it may not be sufficient when dealing with imbalanced datasets or when the costs of misclassification differ across classes. Therefore, we also utilize the F1 score, which is the harmonic mean of precision and recall.

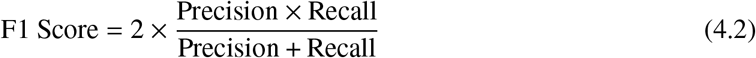

The F1 score balances precision (the proportion of correct positive predictions among all positive predictions) and recall (the proportion of actual positive cases correctly identified), making it particularly valuable for medical diagnostic tasks, where both false positives and false negatives carry significant consequences. These evaluation metrics were reported for each fold in our stratified 5-fold cross-validation setup, while the final performance were determined by averaging the results across all folds.

### 4.4 Results & Analyses

#### 4.4.1 Model Performance

Table 2 presents a comprehensive comparison of the performance based on different AD detection LLM configurations trained on the ADReSSo dataset. Table 2 evaluates the various combinations of LLM (GPT-3.5, GPT-4.0, and GPT4o) input modalities, and prompting strategies. In the modality column, “T” represents the original transcript, “B” denotes the linguistic markers summarised by LLMs, whereas “A” represents the raw audio features. The Audio Model column shows whether Wav2vec is used for audio processing, while the Text Model column indicates whether Bert-Large is used for text processing. The Prompting Strategy column specifies whether the Zero-Shot or Few-Shot (with 10 examples) Learning has been employed. Performance is measured through both Accuracy and F1 score metrics, enabling a balanced assessment of model effectiveness. The results in Table 2 have demonstrated how different combinations of modalities and models affect AD detection performance, with multiple modalities (B+A+T) plus Few-Shot Learning generally achieving better results as compared to single-modality approaches.

**Table 2.**
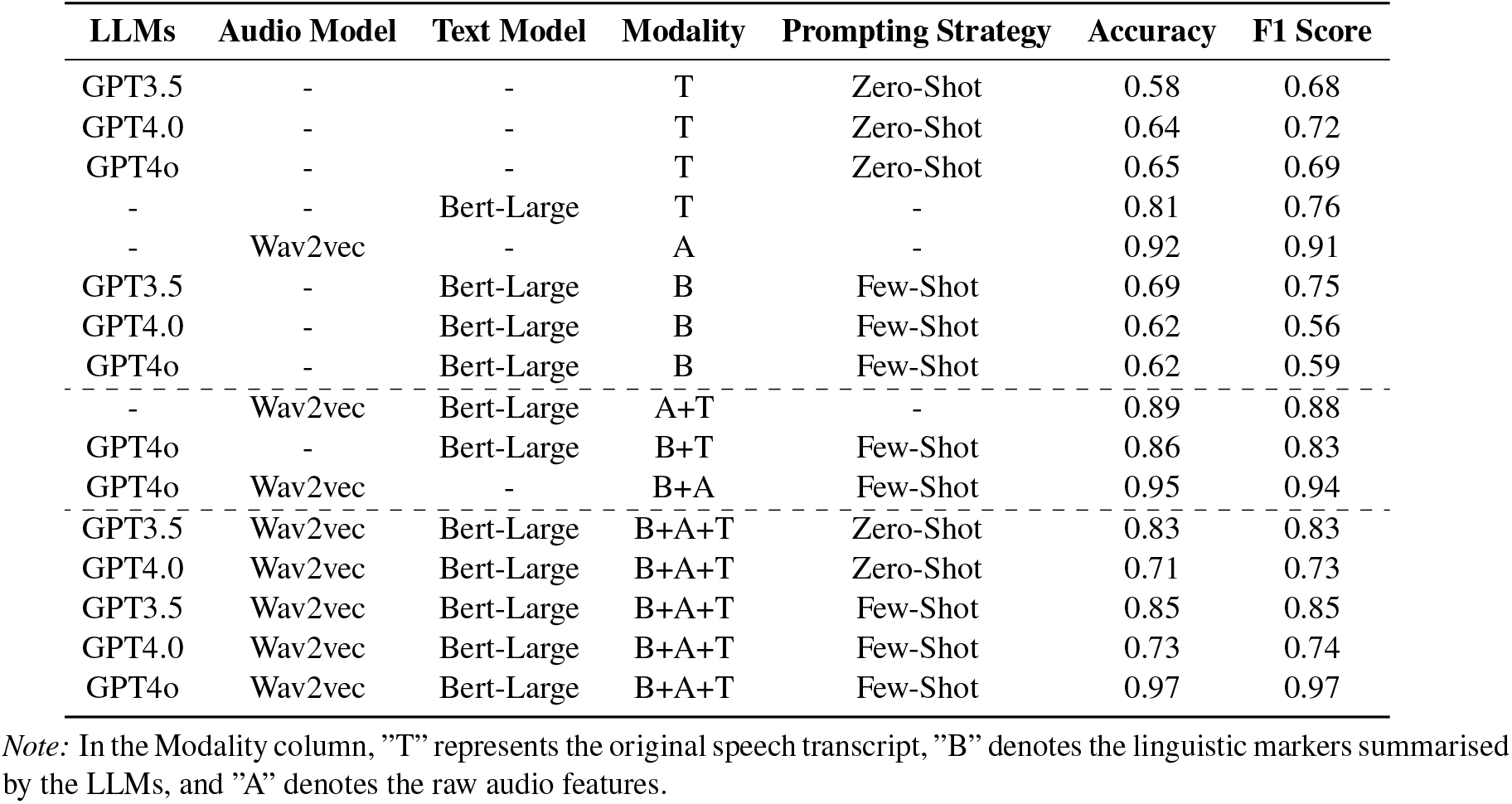
Performance comparison of different methods on the ADReSSo dataset.

Among the single-modality approaches, the audio-based Wav2vec ^34^ model achieves the highest performance (Accuracy = 0.92; F1 score = 0.91), followed by Bert-Large ^4^ using speech transcripts (Accuracy = 0.81; F1 score = 0.76). When LLMs are used alone for diagnosis based on the original transcripts under Zero-Shot Learning, the performance is notably lower, with GPT-3.5 achieving an accuracy of 0.58 (F1 score = 0.68), GPT-4.0 (Accuracy = 0.64; F1 score = 0.72), and GPT4o (Accuracy = 0.65; F1score = 0.69). These results have suggested that these LLMs may not be sufficient for accurate AD detection under the Zero-Shot Learning. The poor performance of the GPT models have raised concerns regarding the feasibility of using the linguistic markers summarised by LLMs to replace the original transcripts. However, the combination of linguistic markers in different modalities have yielded different results. When combining the linguistic markers with the transcripts (B+T), Bert-Large with GPT4o using the Few-Shot Learning (10 shots) has achieved an accuracy of 0.86 (F1 score = 0.83), showing significant improvement over Zero-Shot Learning, which only reaches an accuracy around 0.62. The combination of audio features and linguistic markers (B+A) with GPT4o under the Few-Shot Learning demonstrates impressive performance, achieving an accuracy of 0.95 (F1 score = 0.94).

Most notably, the integration of all three modalities (B+A+T) with GPT4o under Few-Shot learning achieves the best overall performance (Accuracy = 0.97; F1 score = 0.97). This significant improvement over the single-modality approach has suggested that the combination of linguistic markers, acoustic features, and transcript provides the complementary information conducive for more accurate AD detection. These results have revealed that using the acoustic features alone can be effective for AD detection, whilst using the linguistic features alone are not as effective as the acoustic features due to the lack of contextual information. The combination of linguistic features with the original speech transcript or with the audio features, can achieve better results, than using linguistic features alone. The study highlights the potential effectiveness of using linguistic features/markers in combined modalities for AD detection, which are interpretable non-invasive markers highly desirable for clinical diagnosis, when compared to the more invasive biomarkers, such as MRI, or blood biomarkers etc.

### 4.5 Expert-verified Linguistic Markers

To systematically understand the linguistic difference between AD and NC individuals, a linguistic expert has been consulted in order to categorize the various linguistic properties identified by LLMs that distinguish AD from NC cohorts. The expert-categorized linguistic markers are summarized in Table 4. The AD linguistic patterns identified by LLMs are further categorized by the linguistic expert into four major types in Table 3. These linguistic features generally align with what have previously been identified in the literature. ^5,22,23^ First, AD cohorts have displayed problems in lexical recall, resulting in hesitation, repetition, more frequent occurrence of fillers, vague descriptions or even erroneous use of words. In contrast, NC cohorts have displayed little problems with word retrieval and are able to produce fluent speeches. Second, AD cohorts are weak in organizing their thoughts and maintain thematic coherence of their discourse. Their narrations are characterized by unexpected topic switchings as well as incoherent picture descriptions. NC cohorts, in contrast, can organize their thoughts coherently and logically, providing consistent and relevant descriptions. Third, AD cohorts have difficulties in correcting their speech utterances. NC cohorts are able to self-correct their speeches whenever necessary. Lastly, AD cohorts have difficulties in generating grammatical structures while NC cohorts have no problems in generating the right grammatical sentences.

**Table 3.**
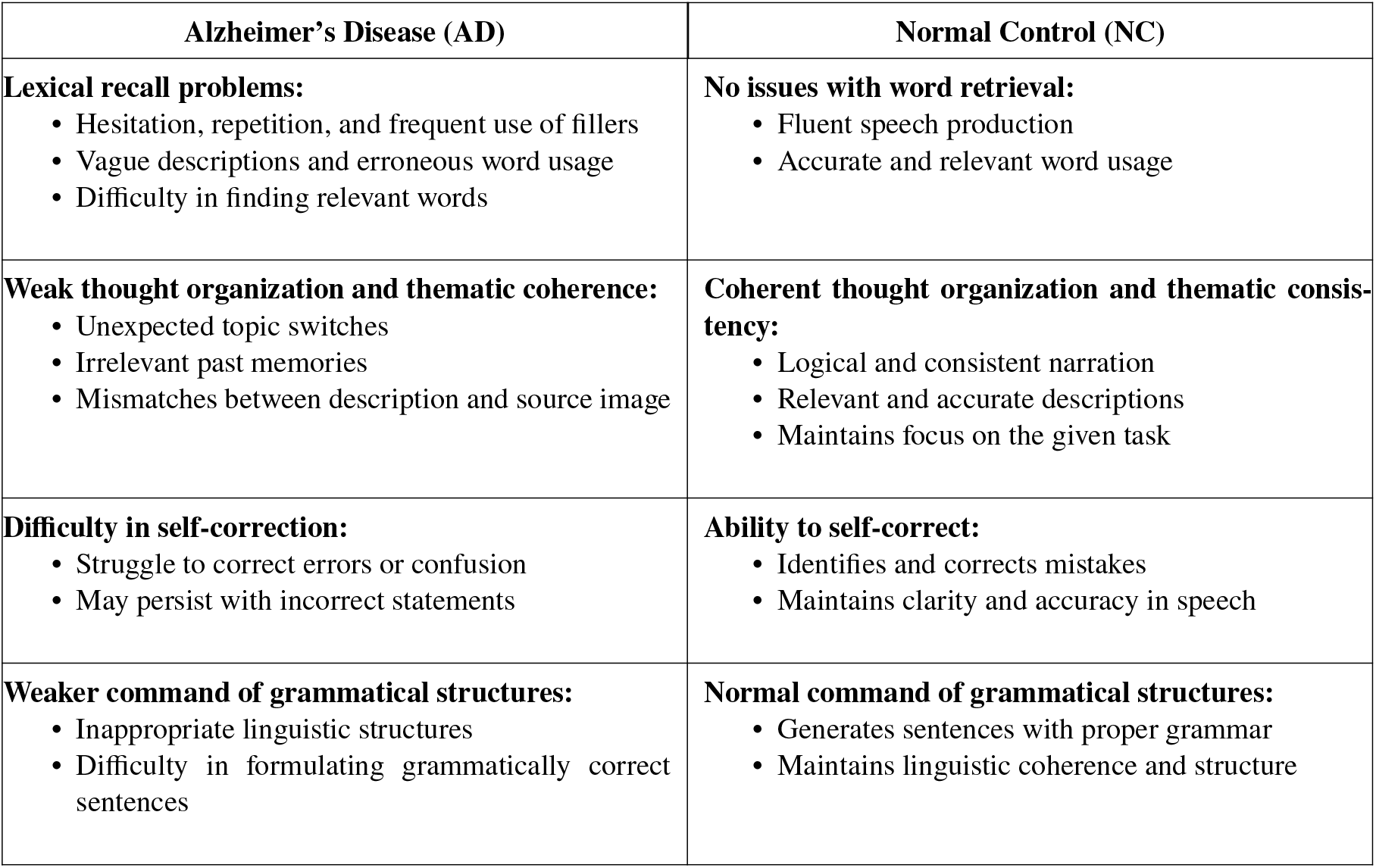
Linguistic features of Alzheimer’s Disease (AD) and Normal Control (NC) cohorts.

**Table 4.**
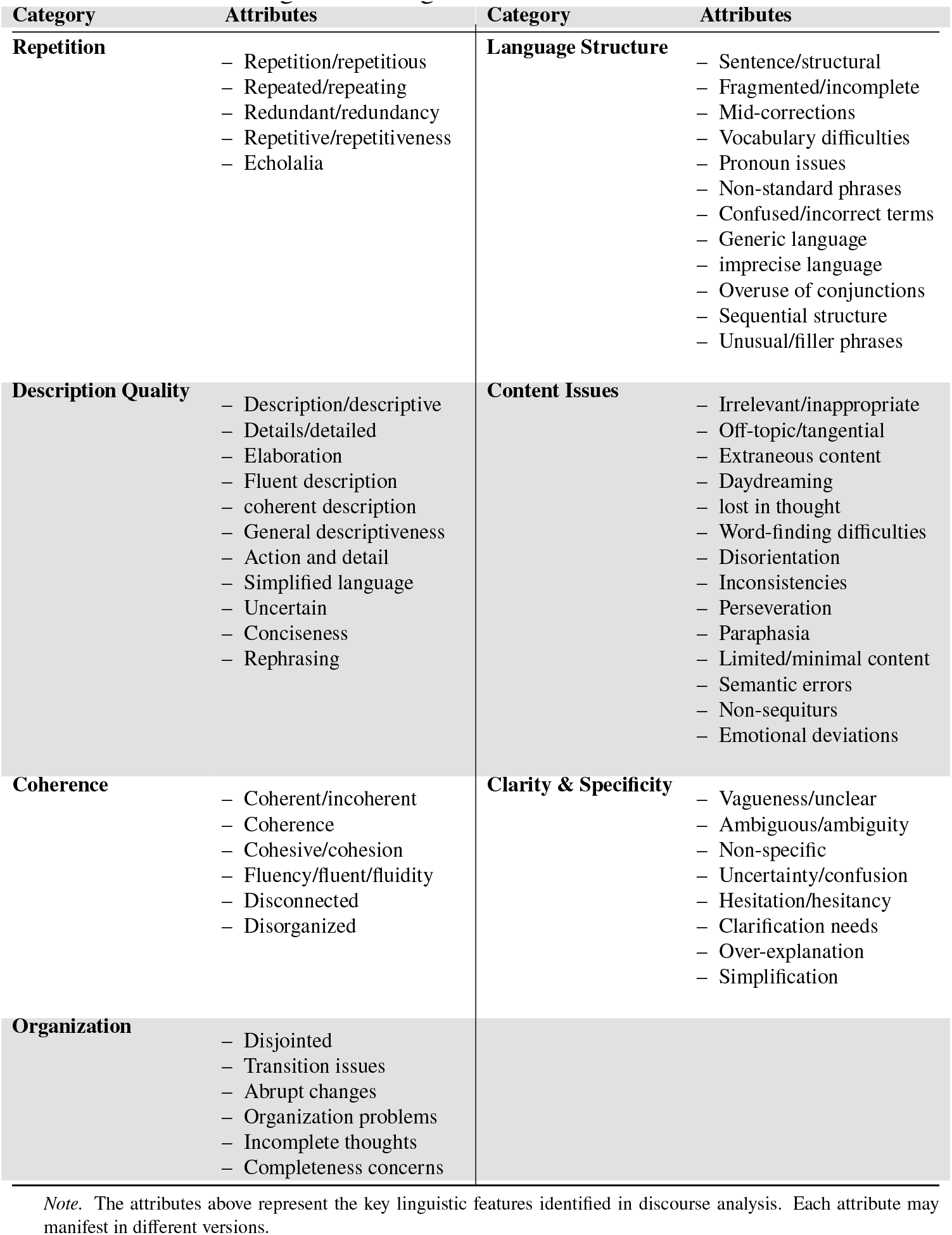
Categories of linguistic markers and their attributes.

### 4.6 LLM-summarised Linguistic Markers

#### 4.6.1 Consistency of LLM-summarised Linguistic Markers Across Different Datasets

To evaluate the stability of LLMs in summarising linguistic patterns from speech samples, we have conducted a comprehensive analysis across multiple datasets. First, we have employed LLMs to identify distinctive language markers that differentiate AD and NC cohorts on three datasets (the Cambridge Dataset, the AdreSSo Dataset, and the Pitt Corpus).Through conducting Similarity Analysis on these LLM-summarised indicators, we have asked the LLMs to classify them into seven categories, based on the shared characteristics for each dataset. Table 4 presents these categories and their corresponding attributes. To determine the frequency of each category, we have computed the semantic similarity between individual indicators and category attributes, which have enabled us to classify the linguistic indicators for each dataset. The frequency distributions across the three AD speech datasets have been shown in Figure 2. These distributions reveal consistent patterns in LLM-summarised linguistic markers, suggesting the robust generalizability of our proposed framework. The most frequently identified linguistic markers have shown remarkable consistency across all three datasets. Repetition has emerged as the predominant marker, with frequencies of 28.9%, 30.1%, and 31.1% in the Cambridge Dataset, the AdreSSo Dataset and the Pitt Corpus, respectively. The high frequency of repetition markers aligns with the previous clinical research, ^30^ which identified repetitiveness as a key predictor of AD. Similarly, Description Quality category corresponds to their finding. The category is the second most frequent across the datasets (FDRD: 27.0%, AdreSSo: 25.0%, Pitt: 29.2%), demonstrating the LLM’s consistency in assessing narrative competence. Language Structure markers have shown minimal variance (0.56) across the three datasets (FDRD: 14.4%, AdreSSo: 13.7%, Pitt: 13.3%), while Clarity and Specificity demonstrated similar stability (FDRD: 13.0%, AdreSSo: 12.5%, Pitt: 11.3%). The stability of Language Structure and Clarity and Specificity markers across datasets is echoed in clinical research ^30^ that identifies grammatical structure and referential specificity as the reliable indicators of cognitive decline. This consistency has also suggested that LLMs can reliably detect syntactic and semantic alterations in speech patterns associated with cognitive decline. The stable hierarchy of markers has suggested that LLMs can reliably prioritize different aspects of linguistic analysis, potentially offering a standardized approach to speech assessment. The consistency across different datasets has demonstrated robust generalization capabilities, thus overcoming the common challenge of inconsistency that LLMs might exhibit when deployed to assist clinician-guided assessment.

**Figure 2.**
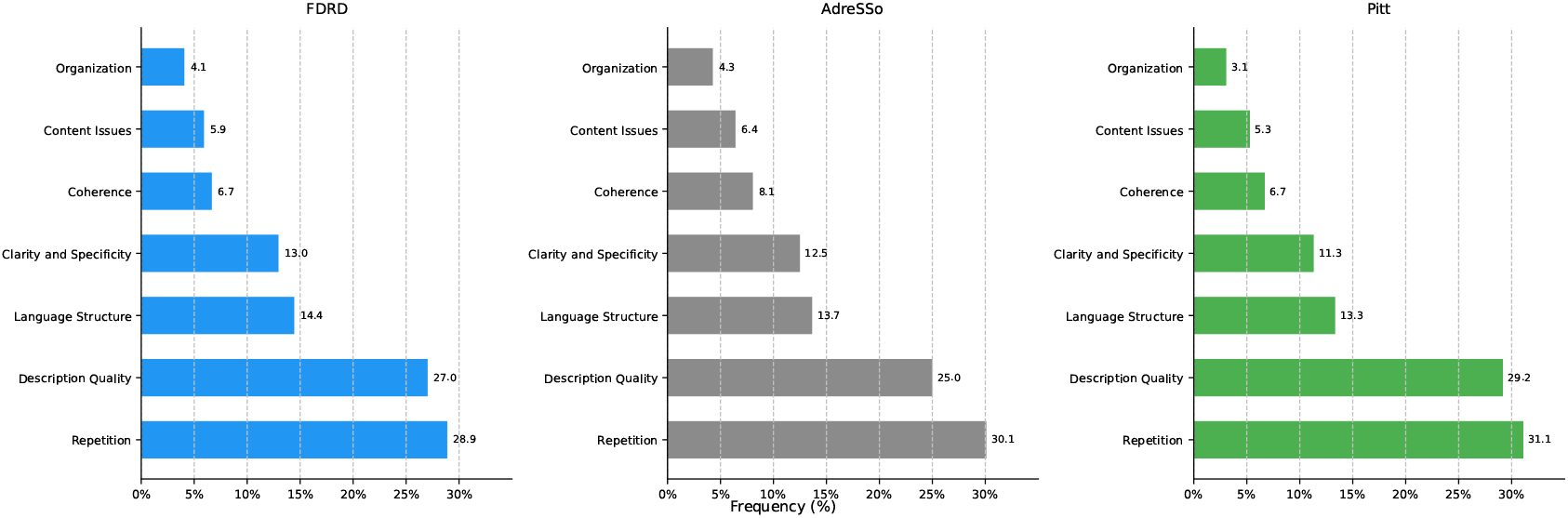
Linguistic marker consistency across three different datasets.

#### 4.6.2 Consistency of LLM-based Responses Across Multiple Iterations

To further ascertain the reliability of LLM-summarised linguistic markers, we examined whether the language patterns identified by LLMs remain consistent across multiple iterations. We repeated the analysis for different number of iterations (3, 5, 7, and 10 times) across three datasets (FDRD, AdreSSo, and Pitt) to determine the optimal number of iterations needed for reliable marker identification. Figure 3 illustrates the distributional patterns of AD linguistic markers by iterations. For each number of iterations, we have analyzed both the variety and frequency of summarised markers, comparing how the patterns stabilize or vary with increased iterations. The visualization reveals that marker distributions has become increasingly stabilized as the number of iterations increases, though in diminishing returns after reaching 5 iterations. This conclusion is supported by quantitative stability metrics showing an average reduction of variability by 47% when the number of iterations increases from 3 to 5, indicating significant stability. Further increase from 7 and 10 iterations has yielded only a marginal improvement (less than 5% reduction in variability). The marker patterns identified across 5 iterations have demonstrated consistent distributions across all three datasets, indicating robust cross-dataset reliability. The threshold of 5 iterations offers a practical guideline for similar applications in AD detection in future. This finding has significant implications for optimizing computational resource while enhancing analytical reliability for consistent clinical decision-making.

**Figure 3.**
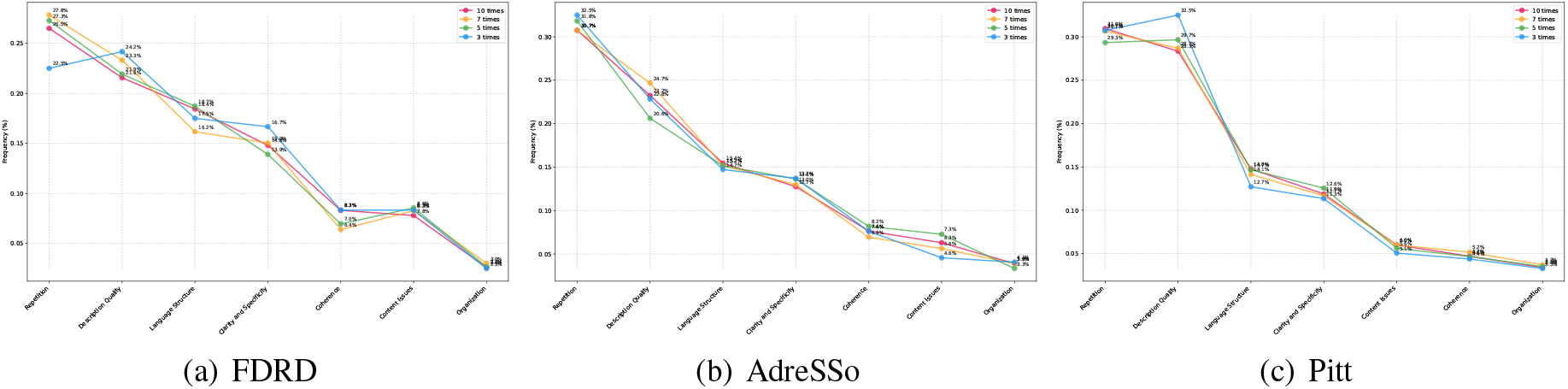
Consistency of linguistic marker across multiple LLM responses.

#### 4.6.3 Frequency of Linguistic Marker Categories across AD vs. NC Cohorts

To compare the distinctive language patterns across the AD and NC cohorts, we have analzyed the language characteristics exhibited by AD and NC across seven key categories. The results in Figure 4 have revealed the linguistic differences between the two groups. Speeches of AD exhibited notably higher frequencies in Description Quality (20.8% vs 18.9%) and Language Structure (16.3% vs 15.1%), as compared to those of NC. Conversely, speeches of NC have demonstrated a higher ability in Clarity and Specificity (14.4% vs. 13.2%), Coherence (9.6% vs. 8.7%), and Organization (4.5% vs. 3.7%). Repetition has comparable occurrence frequency in both the AD and NC cohorts (AD: 29.5%, NC: 30.4%). Figure 4 has displayed minimal between-group variation (AD: 7.7%, NC: 7.2%). These findings suggest a distinctive linguistic pattern, whereas the AD cohorts have shown greater irregularity in descriptive and structural command of language, while the NC cohorts have maintained better command of organizational and coherence-related language acquisition skills. AD cohorts have experienced specific challenge in linguistic production and coherence.

**Figure 4.**
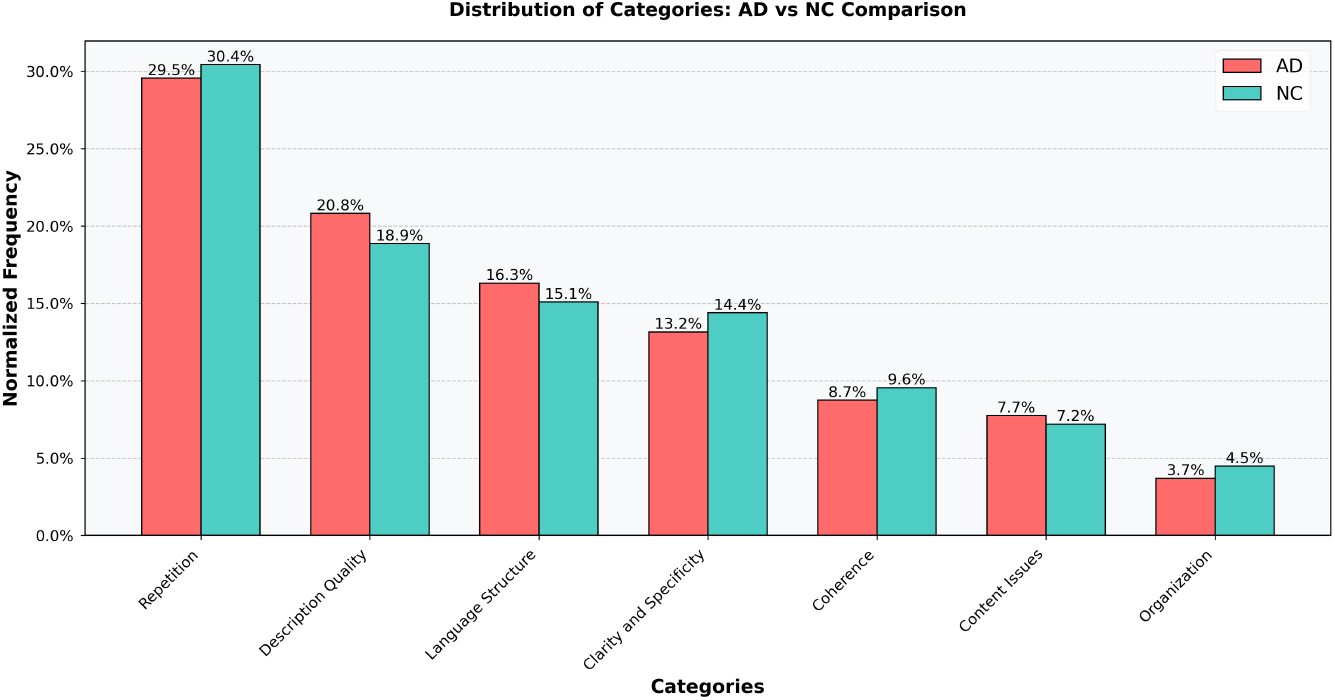
Comparative analysis of linguistic patterns across Alzheimer’s Disease (AD) and Normal Control (NC) cohorts.

## 5 Discussions and the Future Work

In future studies, we aim to refine our search for the distinctive linguistic markers for non-invasive speech-based AD diagnosis. This involves addressing any potential errors or biases arising from LLM-based linguistic marker identification. Linguistic markers are crucial for AD diagnostics for two reasons. First, while the current performance of LLMs in summarising the non-invasive markers may not be highly accurate, it is important to recognize the trade-off between interpretability and accuracy (Rowe, 2024^32^). Linguistic markers summarised by LLMs offer valuable insights into the language patterns and characteristics associated with AD, providing a level of interpretability that is crucial for medical diagnosis and understanding. Although the use of the interpretable linguistic markers may not directly improve accuracy in AD detection, their potential to enhance the interpretability and diagnostic process is invaluable. By leveraging these linguistic markers, healthcare professionals can gain a deeper understanding of the linguistic changes that occur in AD cohorts, enabling the clinicians to make more informed healthcare decisions for the AD patients and provide better patient care.

Second, from the medical and public health perspective, speech-based AD diagnostic offers a great potential as the non-invasive and low-cost solution for AD diagnosis/prognosis, especially when some of these diagnostics may invite scrutiny from a particular stakeholder group (e.g. the patient group) due to time/cost/invasiveness (e.g. MRI/CSF), even though the benefits are huge (Vanderschaeghe et al., 2019; ^31^ Potsteinssen et al., 2021^29^). It has been well recognized that linguistic markers are effective markers for both prediction of AD onset and progression, ^30 2^). Speech-based non-invasive AD diagnosis generally involves relatively simple collection of short speech samples via smartphone apps. They are spontaneous, non-invasive, low-cost and relatively accurate. This means that these apps can be easily deployed for screening and longitudinal speech-based health monitoring. It is in sharp contrast with complicated and expensive traditional AD diagnostics such as MRI, PET imaging, and CSF. As a result, the use of LLMs to identify AD markers is effective, and will substantially benefit diagnosis and prognosis of AD and related clinical/health-based decision making.

The key challenge in summarising accurate and reliable linguistic markers from LLMs is speech data scarcity. With a small amount of speech transcript samples, LLMs may not be able to summarize linguistic markers at high enough accuracy, to make reliable diagnostic decisions and convincing interpretations. To address this challenge, we propose using LLM-based data generation/augmentation techniques to improve the quality of AD speech dataset. Although the LLM-summarised linguistic markers may not be accurate enough for individual AD detection, we can use these markers to personalize AD speech data based on AD-specific linguistic characteristics. By controlling the linguistic patterns and features underlying the AD synthetic speech data, we can create a more diverse and representative dataset covering a broader range of AD-related languistic variations. This approach can help mitigate the limitations of the original dataset (poor in data quality and volume, especially longitudinal data scarcity) and improve the overall performance of AD detection. For instance, we can generate new AD speech samples for facilitating the in-context learning of LLMs. ^25^ By creating new speech examples that mimic the linguistic patterns and characteristics of AD, we can provide LLMs with larger and more diverse (e.g. longitudinal) datasets. This new AD speech samples can help LLMs learn and extract more accurate linguistic markers and improve their diagnostic performance. Moreover, we can also generate speech transcript samples for supervised multimodal model learning and training. In addition to enhancing the performance of LLMs, the new AD speech data can also be used to train the multimodal model by combining linguistic markers with other modalities. By generating a larger dataset with pre-defined characteristics, we can explore a wider sample space and potentially improve the training performance of multimodal model training.

## 6 Conclusion

In conclusion, this study demonstrates the potential of using LLMs to extract interpretable linguistic markers for speech-based non-invasive AD detection. By leveraging the in-context Few-Shot and Zero-Shot learning and prompting strategies, we show that LLMs can identify high-level linguistic patterns discriminating AD from NC. Our designed prompting strategies facilitate LLMs by providing interpretation and assessment of the summarised markers, showcasing the strength, reliability, and relevance of LLMs for AD classification. Furthermore, we propose a novel framework that incorporates the linguistic markers summarised by LLMs into a small supervised-learning model. Our comprehensive analyses have shown that while acoustic features alone are highly effective for AD detection, linguistic markers summarised by LLMs in isolation lack the sufficient contexts for accurate linguistic-based classification. When these linguistic markers are integrated with either acoustic features or original speech transcripts through Few-Shot learning approaches, they can significantly enhance AD detection performance, achieving 97% accuracy (maximum) for AD classification. In future studies, we should focus on improving the accuracy of LLM-summarised markers while maintaining interpretability. Investigating speech data generation/augmentation techniques is also a promising direction. Overall, this study takes an important step in harnessing the power of LLMs for interpretable speech-based AD detection, paving the way for future research. By combining the strength of LLMs with supervised learning, we can work towards developing more accurate, interpretable, and trustworthy LLM-based AD detection systems to assist the healthcare professionals for early diagnosis and therapeutic care for the AD patients, helping LLMs fulfil the desirable and meaningful purpose of safeguarding the well-being of our society, by facilitating sound LLM-based clinical decision-making.

## Data Availability

All data produced are available online at https://dementia.talkbank.org/ADReSSo-2021/

## 7 Appendix-A

**Table 5.**
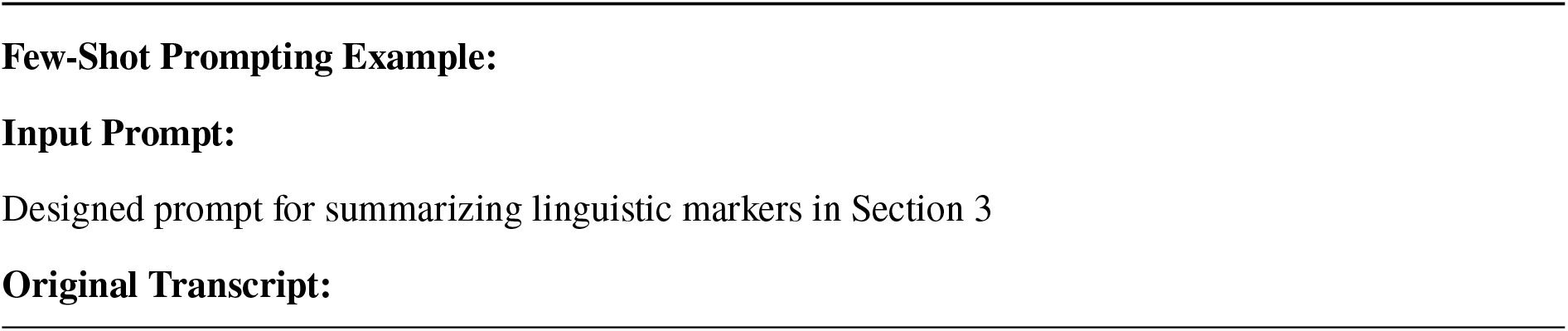
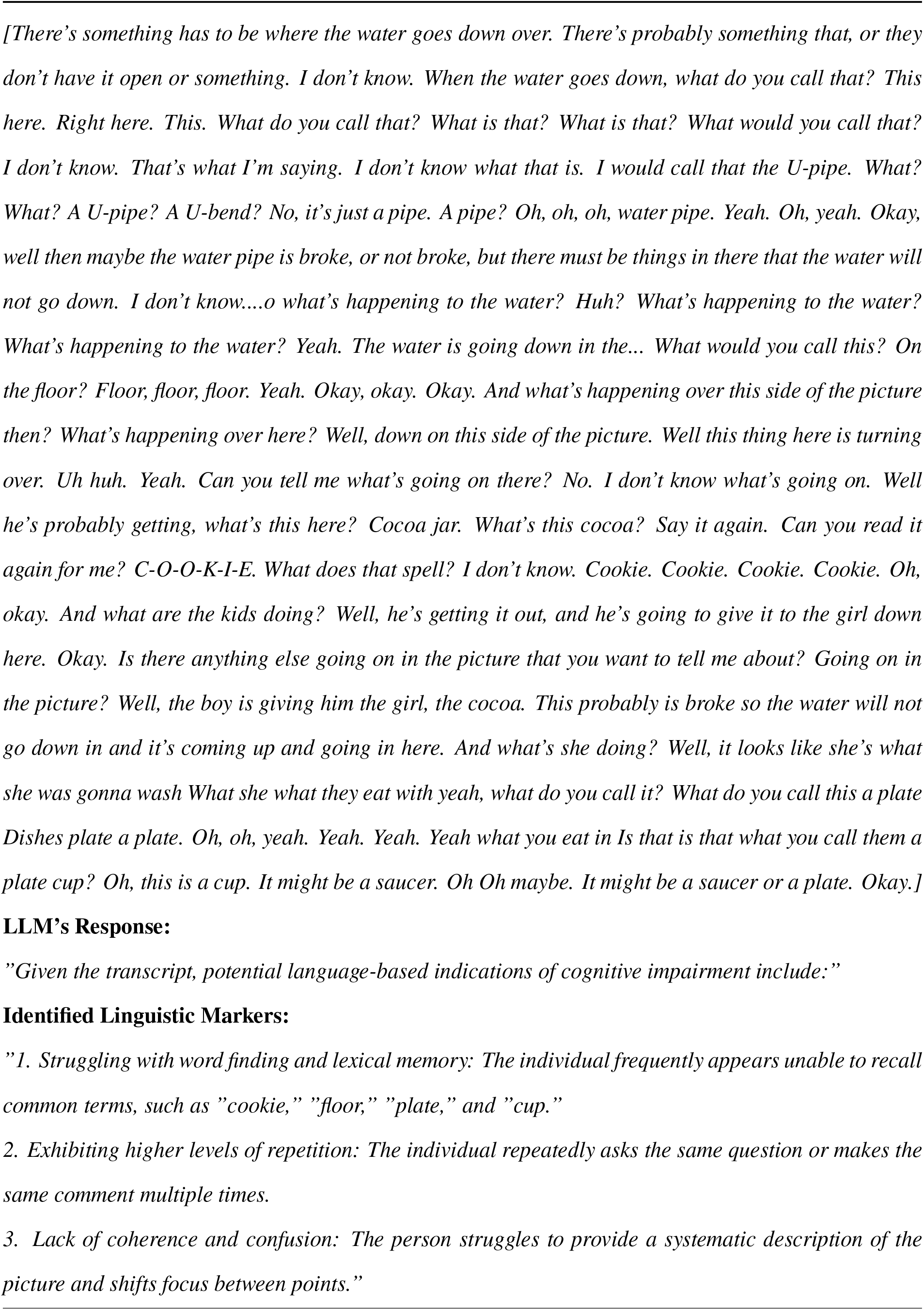
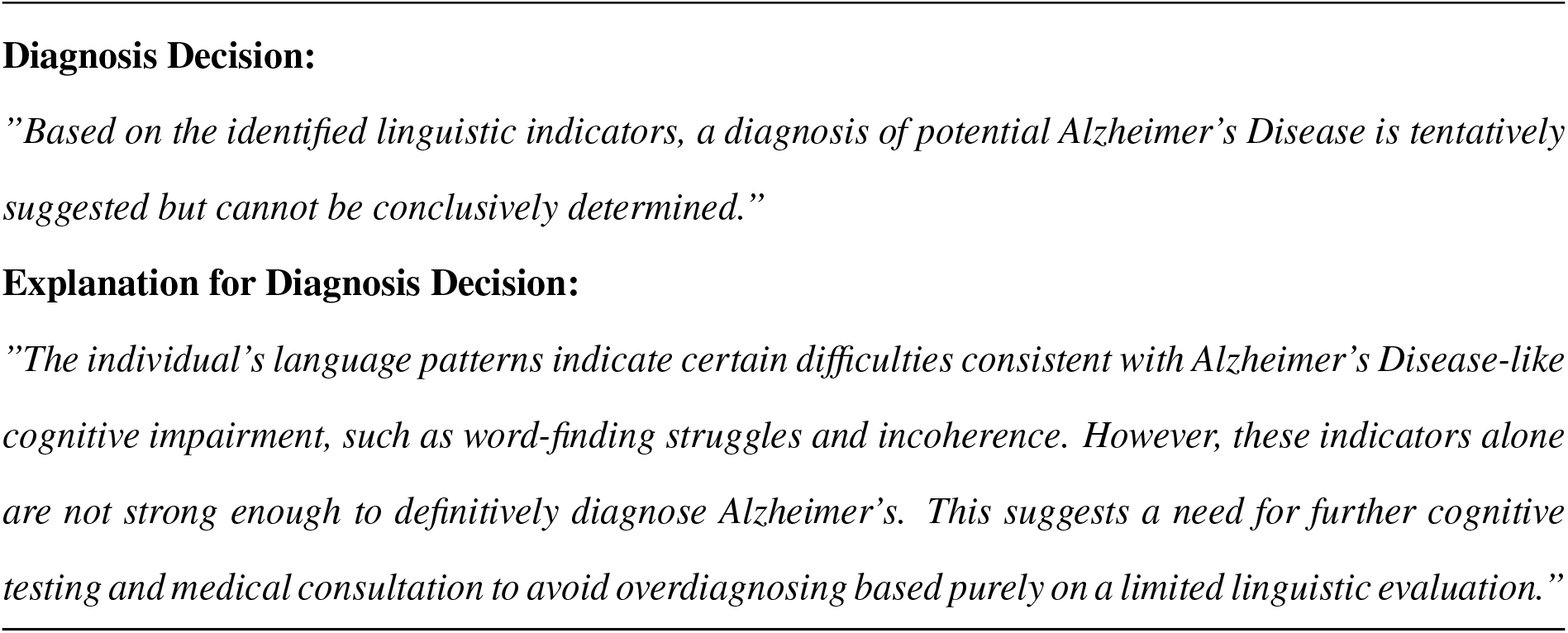
Example of using LLMs to identify linguistic markers.

## 8 Appendix-B

**Table 6.**
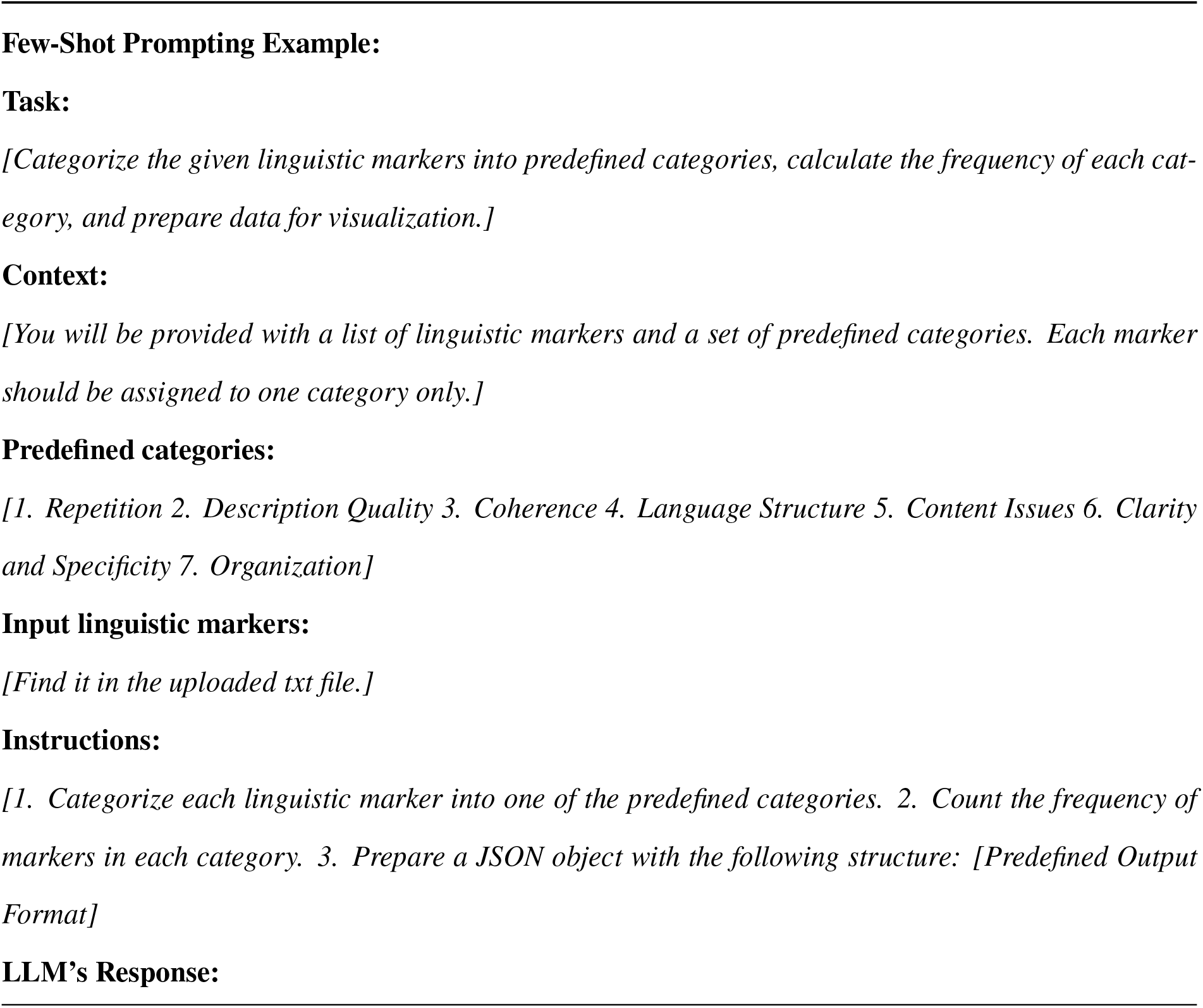
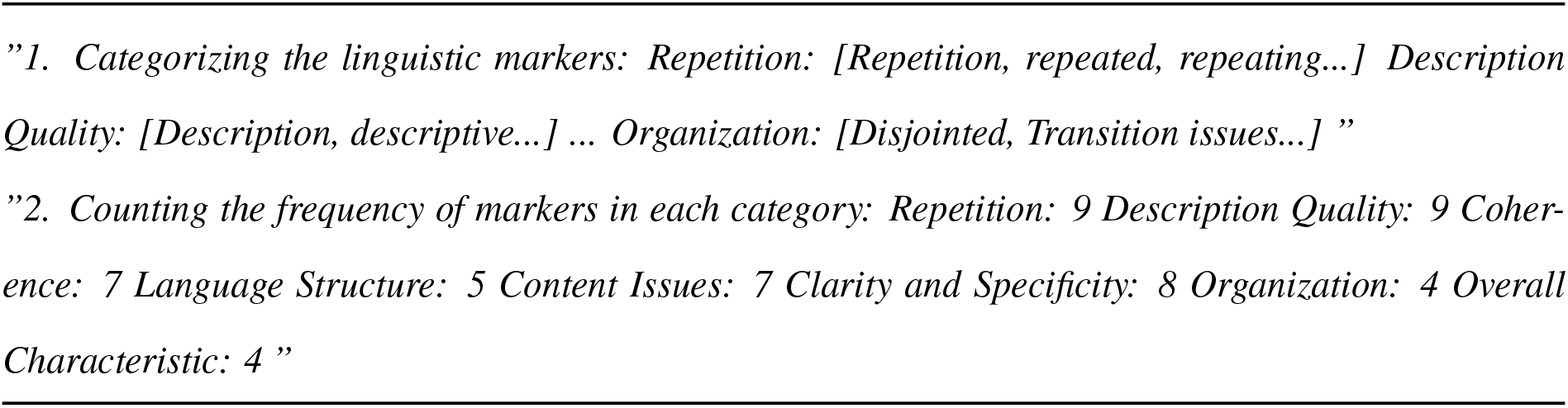
Example of LLMs categorizing linguistic markers.

## Acknowledgements

We gratefully acknowledge Yang Han for the valuable discussions and insights that greatly benefited our work. We would also like to thank Prof. James Rowe and Prof. David Rubinstein, the University of Cambridge, for their helpful insights and suggestions for this study.

## Author Contributions

Conceptualization,V.O.K.L.,J.C.K.L; Methodology,T.Y.M.,J.C.K.L., V.O.K.L; Formal Analysis,T.Y.M, J.C.K.L., L.Y.L.C; Data Curation, T.Y.M.; Writing—Original Draft, T.Y.M., Writing—Review and Editing, J.C.K.L. and V.O.K.L; Supervision, J.C.K.L. and V.O.K.L.; Funding Acquisition, J.C.K.L. and V.O.K.L.

## Footnotes

1 The FDRD dataset used in this study was derived from Cambridge Centre for Frontotemporal Dementia and Related Disorders. IRB approval has been obtained by our collaborator, Prof. James Rowe, from the University of Cambridge.

